# Clinical Performance of LucentAD Complete: A Multi-Analyte Algorithmic Blood Test for Detecting Brain Amyloid with a Scalable, Multiplexed Digital Immunoassay

**DOI:** 10.1101/2025.03.31.25324984

**Authors:** David Wilson, Karen Copeland, Ann-Jeanette Vasko, Lyndal Hesterberg, Meenakshi Khare, Michele Wolfe, Patrick Sheehy, Inge M.W. Verberk, Mike Miller, Charlotte E. Teunissen

**Author notes:** Address correspondence to: David H Wilson, PhD.

## Abstract

**INTRODUCTION:** To address an urgent need for a scalable, accurate blood test for brain amyloid pathology that provides a conclusive result for the greatest number of patients, we developed a multi-analyte algorithmic test combining p-Tau 217 with four other biomarkers.

**METHODS:** Multiplexed digital immunoassays measured p-Tau 217, Aβ42/40, GFAP, and NfL in 730 individuals to establish an algorithm with cutoffs, then in 1,082 individuals with cognitive symptoms from three independent cohorts to identify brain amyloid pathology.

**RESULTS:** The test’s algorithmic risk score AUC was 0.92, yielding 90% agreement with amyloid PET and CSF. Positive predictive value was 92% at 55% amyloid prevalence. The multi-marker algorithm reduced the intermediate zone 3-fold to 12% vs. p-Tau 217 alone. Diagnostic performance was similar by race, ethnicity, sex, age, and apoE4 status.

**DISCUSSION:** The LucentAD Complete multi-analyte blood test demonstrated high clinical validity for brain amyloid pathology detection while substantially reducing inconclusive intermediate results.

## 1. BACKGROUND

The advent of new Alzheimer’s therapies underscores the urgent need for improved early diagnostic tools. While clinical diagnosis has historically been the standard, biomarkers, particularly amyloid and phosphorylated tau, are increasingly crucial for an etiological diagnosis and eligibility for amyloid targeting therapies. Traditionally, these biomarkers required costly and invasive PET or CSF assessments. However, recent rapid advancements in blood-based biomarker tests offer non-invasive alternatives for amyloid pathology assessment, promising expanded access to diagnosis through simpler, more efficient procedures. Among blood-based biomarkers, tau phosphorylated at threonine 217 (p-Tau 217) has emerged as the most accurate single biomarker for detecting amyloid pathology in Alzheimer’s disease (AD), with the Alzheimer’s Association (AA) recommending it as the only blood-based biomarker comparable to FDA-cleared CSF tests for diagnostic use, when analyzed with high performance technologies [1]. Recognizing overlap between amyloid-positive and -negative assay signals, the AA and others advocate for a two-cutoff test design, creating an intermediate risk zone [2]. To maximize clinical utility, the AA and Global CEO Initiative (CEOi) recommend an amyloid classification accuracy of ≥90% and an intermediate zone of ≤20% [3]. Accurately classifying borderline cases within this intermediate zone remains a key challenge.

Beyond p-tau 217, several other established plasma biomarkers are relevant to detecting the presence of amyloid pathology. These biomarkers, including the amyloid β 1-42/1-40 ratio (Aβ42/Aβ40), glial fibrillary acidic protein (GFAP), and neurofilament light chain (NfL), reflect amyloid pathology directly or detect the presence of AD-associated disease pathways. Aβ42/Aβ40 directly reflects amyloid plaque development [4], GFAP indicates astrocytic activation linked to amyloid pathogenesis [5], and NfL signals neuroaxonal damage in neurodegenerative diseases like AD [6]. While these biomarkers can individually predict AD and their predictive power is enhanced in combination [7], they are less accurate than p-Tau 217 alone [8] and combining them with p-Tau 217 has shown some, but minimal overall accuracy improvement [9, 10, 11]. In this multi-cohort clinical validation study, we discovered that the combination of these five biomarkers dramatically improved amyloid classification of borderline intermediate cases compared to p-Tau 217 alone, resulting in a substantial reduction of the intermediate zone while preserving high overall test accuracy. Thus, the approach combines the accuracy of p-Tau 217 for cases with low and high amyloid burden, with additional amyloid classification ability for borderline cases through the addition of Aβ42/Aβ40, GFAP, and NfL. These four augmentative biomarkers are combined in a single multiplexed Simoa digital immunoassay (N4PE), which is combined with the LucentAD p-Tau 217 immunoassay to provide accurate amyloid classification results for more patients than possible with p-Tau 217 alone. This report describes the test, and its clinical performance based on a multi-cohort clinical validation study utilizing pre-established diagnostic cutoffs. The test output is a logistic probability (risk) score interpreted as low, intermediate, or high risk for amyloid pathology. These validation data support availability of the test under CLIA (as “LucentAD Complete”) for clinical use to provide accurate results to the greatest number of patients with a simple, scalable immunoassay format.

## 2. METHODS

### 2.1 Cohorts and reference methods

#### 2.1.1 Amsterdam Dementia Cohort

The Alzheimer Center Amsterdam (ADC) cohort comprised patients referred for cognitive evaluation by general practitioners or specialists. All patients underwent a standardized, multidisciplinary work-up, including neurological history and examination, vital function assessment, informant history, dementia nurse consultation, neuropsychological testing, brain MRI, EEG, standard laboratory tests, and either lumbar puncture for CSF biomarker analysis or amyloid PET imaging [12, 13]. Diagnoses were determined via multidisciplinary consensus, with Alzheimer’s dementia requiring abnormal CSF biomarkers or positive amyloid PET [14, 15]. Amyloid PET scans utilized [18F]Florbetaben or [18F]Florbetapir, with positivity defined by visual assessment of neocortical fibrillary amyloid by a nuclear medicine physician. CSF amyloid positivity was determined using Roche Elecsys p-tau 181/Aβ42 assays (cut-off 0.02) [16] or Fujirebio Innotest p-tau 181/Aβ42 ELISAs (cut-off 0.06) [17]. As the intended use population is objectively impaired patients, MCI (n=403) and AD (n=336) cases constituted 68% of the training and 25% of the validation sample sets (demographic characteristics previously reported [17]). Demographic characteristics of the sub cohort used for validation (n=274) are shown in Table 1. For analysis of amyloid detection accuracy in mixed pathology cases, we also examined subgroups of symptomatic individuals diagnosed with vascular dementia (VaD, n=60), frontotemporal dementia (FTD, n=220), and dementia with Lewy bodies (DLB, n=182), some of whom also had amyloid pathology. Demographic data for these subgroups are provided in the supplemental materials.

**TABLE 1.** Demographic and clinical characteristics.

|  | <u>BH (N = 271)</u> | <u>ADC (N = 274)</u> | <u>ADNI (N = 537)</u> | <u>All (N= 1082)</u> |
| --- | --- | --- | --- | --- |
| **Age** |  |  |  |  |
| Mean (SD) | 73.4 (6.8) | 64.9 (7.7) | 70.6 (7.1) | 69.9 (7.8) |
| Range | 60-85 | 44-83 | 55-86 | 44-86 |
| **Sex** |  |  |  |  |
| F | 148 (54.6%) | 130 (47.4%) | 241 (44.9%) | 519 (48.0%) |
| M | 123 (45.4%) | 144 (52.6%) | 296 (55.1%) | 563 (52.0%) |
| **Race** |  |  |  |  |
| Native American or Alaskan | 0 (0.0%) | 0 (0.0%) | 3 (0.6%) | 3 (0.3%) |
| Asian | 8 (3.0%) | 0 (0.0%) | 7 (1.3%) | 15 (1.4%) |
| Black or African American | 30 (11.1%) | 0 (0.0%) | 17 (3.2%) | 47 (4.3%) |
| More than one race | 0 (0.0%) | 0 (0.0%) | 4 (0.7%) | 4 (0.4%) |
| Unknown | 4 (1.5%) | 0 (0.0%) | 0 (0.0%) | 4 (0.4%) |
| White | 229 (84.5%) | 274 (100.0%) | 506 (94.2%) | 1,009 (93.3%) |
| **Ethnicity** |  |  |  |  |
| Hispanic or Latino | 31 (11.4%) | 0 (0.0%) | 27 (5.0%) | 58 (5.4%) |
| Not Hispanic or Latino | 239 (88.2%) | 274 (100.0%) | 510 (95.0%) | 1,023 (94.5%) |
| Not Reported | 1 (0.4%) | 0 (0.0%) | 0 (0.0%) | 1 (0.1%) |
| **MMSE** |  |  |  |  |
| Mean (SD) | 25.4 (2.9) | 24.3 (4.9) | 26.5 (4.0) | 25.6 (4.1) |
| Range | 16-30 | 2-30 | 7-30 | 2-30 |
| Missing | 0 (0.0%) | 4 (1.5%) | 118 (22.0%) | 122 (11.3%) |
| **Diagnostic Group** |  |  |  |  |
| AD | 0 (0.0%) | 96 (35.0%) | 93 (17.3%) | 189 (17.5%) |
| Mild AD | 122 (45.0%) | 0 (0.0%) | 0 (0.0%) | 122 (11.3%) |
| MCI | 149 (55.0%) | 178 (65.0%) | 444 (82.7%) | 771 (71.3%) |
| **ApoE** |  |  |  |  |
| ε2ε3 | 28 (10.3%) | 11 (4.0%) | 42 (7.8%) | 81 (7.6%) |
| ε2ε4 | 6 (2.2%) | 4 (1.5%) | 10 (1.9%) | 20 (1.9%) |
| ε3ε3 | 135 (49.8%) | 91 (33.2%) | 298 (55.5%) | 524 (48.9%) |
| ε3ε4 | 87 (32.1%) | 108 (39.4%) | 156 (29.1%) | 351 (32.7%) |
| ε4ε4 | 15 (5.5%) | 50 (18.2%) | 31 (5.8%) | 96 (9.0%) |
| ε4 carriers | 108 (39.9%) | 162 (59.1%) | 197 (36.7%) | 467 (43.6%) |
| Missing | 0 (0.0%) | 10 (3.6%) | 0 (0.0%) | 10 (0.9%) |
| **Amyloid Status** |  |  |  |  |
| CSF | 0 (0.0%) | 263 (96.0%) | 0 (0.0%) | 263 (24.3%) |
| PET | 271 (100.0%) | 11 (4.0%) | 537 (100.0%) | 819 (75.7%) |
| Amyloid Prevalence | 45.4% | 70.8% | 52.3% | 55.4% |
| Amyloid Risk Score (SD) | 59.6 (32.8) | 64.2 (32.8) | 71.7 (31.9) | 61.8 (32.4) |
| Amyloid Risk Score, range | 0.00-1.00 | 11.3-1.00 | 12.4-1.00 | 0.00-1.00 |

Notes: Validation cohorts. Abbreviations: BH, Bio-Hermes; ADC, Amsterdam Dementia Cohort; ADNI, Alzheimer's Disease Neuroimaging Initiative; MCI, mild cognitive impairment; PET, positron emission tomography.

#### 2.1.2 Bio-Hermes Cohort

From April 2021 to November 2022, 17 clinical trial sites recruited community-based participants for the Bio-Hermes cohort, aiming to enrich ethnic/racial diversity. Participants, meeting established inclusion criteria [18], were categorized as cognitively unimpaired, MCI, or mild AD. MCI participants had a documented MCI diagnosis (NIA-AA criteria [19]) or met screening criteria: MMSE 24-30, RAVLT-delayed recall ≥1 SD below age-adjusted mean, and minimal functional impairment (FAQ). Mild AD participants had a probable AD diagnosis (NIA-AA criteria [20]) or met screening criteria: MMSE 20-24, RAVLT-delayed recall ≥1 SD below age-adjusted mean, and evidence of functional decline (FAQ). All participants underwent [18F]Florbetapir amyloid PET scans, interpreted centrally by IXICO Technologies Inc. Underrepresented groups (Hispanic and non-Hispanic Black) comprised 27.8% of the symptomatic sub-cohort (MCI and mild AD). Consistent with the intended use population, MCI (n=285) and mild AD (n=221) cases constituted 32% of the training and 25% of the validation sample sets. Details of these subgroups have been previously reported [18]. Demographic characteristics of the sub cohort used for validation (n=271) are shown in Table 1.

#### 2.1.3 ADNI Cohort

The ADNI cohort, derived from the ADNI database (adni.loni.usc.edu) [20], comprised participants from a FNIH Biomarker Consortium study examining longitudinal trajectories of blood-based biomarkers in relation to amyloid PET [21]. ADNI, initiated in 2003, aims to assess the progression of MCI and early AD using multimodal biomarkers. For this validation study, participants with plasma samples collected within 6 months of amyloid PET at three distinct time points were selected. From 406 subjects with 1231 samples (2-5 time points), 1010 had amyloid status and all five biomarker assays. Exclusion of samples outside the intended use population yielded 537 samples from 236 individuals, spanning up to 180 months. A subset of 107-115 individuals with contiguous baseline, 24-month, and 48-month time points was analyzed separately to assess test result consistency over time. The ADNI cohort was not utilized in training and constituted 50% of the validation sample set. Demographic characteristics of the ADNI cohort are shown in Table 1.

#### 2.1.4 Validation vs. training cohorts

p-tau 217 clinical thresholds were established using approximately 50% of the ADC cohort 2 and a randomized subset of the Bio-Hermes training and validation cohorts, as previously described [22]. All p-tau 217 testing for this threshold determination was performed at the Quanterix CLIA laboratory using a single reagent/calibrator lot. Multi-analyte algorithm thresholds were derived from N4PE data on ADC cohort 1 (n=495, Neurochemistry Laboratory Amsterdam) and the Bio-Hermes training sub-cohort (n=235, Quanterix CLIA laboratory). To account for different reagent lots used between laboratories, a crossover set of 100 samples was tested at both sites. Bridging of individual biomarkers was made using Passing-Bablock regression, with an average bridging adjustment of 15-22% depending on the biomarker. The validation of the multi-analyte thresholds was conducted exclusively at the Quanterix CLIA laboratory, using ADC cohort 2 (n=274) combined with the Bio-Hermes validation sub-cohort (n=271), as well as the independent ADNI longitudinal cohort (n=537).

### 2.2 Plasma sample analysis

#### 2.2.1 Instrumentation

All assay testing was performed on the Simoa HD-X instrument, a fully automated digital immunoassay analyzer utilizing Simoa technology for isolation and counting of single molecules. Details of the instrument and its principles are given elsewhere [23].

#### 2.2.2 Assay principle and protocol

Simoa technology, a digitized bead-based ELISA, achieves attomolar sensitivity through single-molecule detection within 40-femtoliter microwells [24]. By confining fluorescent reporter molecule diffusion, single enzyme labels generate detectable signals within 30 seconds. Arrays of 216,000 wells enable rapid, simultaneous counting, resulting in assay completion within 45-60 minutes.

The Simoa p-tau 217 assay, a 3-step sandwich immunoassay, involves capture, sandwich formation with biotinylated detector antibodies, and labeling with a streptavidin-β-galactosidase conjugate [22]. Following magnetic bead collection and washing at each step, beads are resuspended in resorufin β-D-galactopyranoside substrate. Digital processing occurs upon bead transfer to the Simoa array disc, where captured p-tau 217 leads to substrate hydrolysis and fluorescence. p-tau 217 concentration is determined via 4-parameter logistic curve interpolation, with assay completion in approximately one hour.

The commercially available Simoa Neurology 4-plex E Kit (N4PE) simultaneously measures Aβ40, Aβ42, GFAP, and NfL using a 2-step digital immunoassay. Capture beads and detector antibodies are combined, with analyte-specific beads pre-coated with distinct fluorescent dyes. Following substrate resuspension and array transfer, bandpass filters identify analyte-specific beads. Concentrations are determined through 4-parameter logistic regression fitting, with a processing time of approximately one hour.

#### 2.2.3 Sample collection and testing

K2EDTA plasma was collected via venipuncture for all three cohorts. For the Amsterdam Dementia Cohort (ADC), samples were centrifuged within 2 hours (1800xg, 10 min, room temperature), aliquoted (≤0.5mL), and stored at -80°C until transfer/shipment to either the Neurochemistry Laboratory Amsterdam or Quanterix. In Amsterdam, samples were thawed, centrifuged (10,000xg, 10 min), and analyzed in singlicate for Aβ40, Aβ42, GFAP, and NfL using the Simoa N4PE kit on the Simoa HDx analyzer, with calibration and quality controls in duplicate. Inter-assay coefficients of variation were <15% [17]. For the Bio-Hermes cohort, 2.0mL whole blood was transferred to conical tubes and centrifuged (1500xg, ≥15 min, room temperature), with plasma aliquoted and frozen at -80°C within 4 hours. For the ADNI cohort, plasma was centrifuged within 1 hour (1500xg, 15 min, room temperature), aliquoted, and frozen at - 80°C. For testing at Quanterix, samples were thawed (60 min, room temperature), centrifuged (10,000xg, 10 min), and analyzed), with calibrators, controls, and quality controls according to Quanterix SOPs.

Quality control samples, included in all p-Tau 217 and N4PE runs, demonstrated acceptable precision, consistent with previous reports [17, 22]. The precision of the composite multi-analyte Amyloid Risk Score, incorporating all five biomarkers, was estimated by evaluating inter-assay imprecision from a multi-day precision study for each assay. The expected composite score imprecision was then modeled from the combined biomarkers, as described in the supplemental materials. The overall expected mean inter-day imprecision (%CV) of the composite Amyloid Risk Score was 7.33% (95% CI: 6.87-7.80%).

### 2.3 Statistical methods

All data analysis was performed using JMP Pro 18 (JMP Statistical Discovery LLC, Cary, NC). Discovery analyses, employing multiple modeling techniques, graphical data interrogations, and performance metrics, resulted in the LucentAD Complete test configuration, consisting of a locked algorithm and thresholds. Test performance was subsequently validated in three independent cohorts, both individually and combined. Performance metrics, including 95% confidence intervals, are reported, with specific mention of instances where indeterminate zone results were excluded.

The test algorithm builds upon established thresholds for the p-Tau 217 assay. Initially, a logistic regression model was constructed using all five biomarkers across the training dataset. A separate logistic regression model transformed p-Tau 217 levels into a model score. These two models were then combined using a decision tree based on the p-Tau 217 thresholds. Thresholds for the resulting multi-analyte model score were determined by fitting best-fit distributions to the negative and positive training samples. Specifically, the upper threshold was set at the 93rd percentile of a sinh-arcsinh distribution fit to the negative samples (corresponding to a 7% false positive rate), and the lower threshold was set at the 7th percentile of a beta distribution fit to the positive samples (corresponding to a 7% false negative rate). These 7% false negative and false positive rates were selected to establish robust thresholds, aiming for 10% false negative and false positive rates in independent validation cohorts.

## 3. RESULTS

### 3.1 Demographic and clinical characteristics

Demographic and clinical characteristics of the three validation cohorts, and the combined validation set stratified by amyloid status, are detailed in Table 1 and Table S1, respectively. The combined validation cohort had a mean age of 69.9 years (SD 7.8, range 44-86), with 48.0% female representation. Mean age varied across cohorts: ADC, 64.9 years (SD 7.7, range 44-83); Bio-Hermes, 73.4 years (SD 6.8, range 60-85); and ADNI, 70.6 years (SD 7.1, range 55-86). The validation set was predominantly White (93.3%), with Bio-Hermes contributing the majority of underrepresented minorities (25.5% within-cohort proportion). All participants were symptomatic, with diagnoses of MCI (71.3%), mild AD (11.3%), or AD (17.5%). 43.6% were ApoE ε4 carriers. Amyloid prevalence varied significantly across cohorts and clinical subgroups: ADC, 67.3% in MCI and >99% in AD dementia (CSF positivity required for selection); Bio-Hermes, 34.7% in MCI and 61.5% in dementia (PET confirmed); and ADNI, 45.1% in MCI and 88.2% in dementia (PET confirmed). The combined validation set had an overall amyloid prevalence of 55.4%. These variations likely resulted from differences in the clinical determination of diagnostic categories (MCI, AD dementia), CSF requirement for diagnosis in ADC, and Bio-Hermes diagnoses preceding PET testing.

### 3.2 Analyte measurement in plasma samples

K2EDTA plasma samples from ADC (n=769), Bio-Hermes (n=506), and ADNI (n=537) were analyzed for p-tau 217, Aβ42/Aβ40, GFAP, and NfL, with results compared to reference amyloid status (CSF or PET). Figure 1 displays biomarker results stratified by amyloid status. All samples were above the assay LoD, and 99.5% of p-tau 217 samples exceeded the LLoQ, ensuring quantifiable data. All other assays yielded 100% LLoQ-exceeding samples. Median p-tau 217 concentration was 3.14-fold higher in amyloid-positive participants (negative: 0.035 pg/mL, IQR 0.02; positive: 0.11 pg/mL, IQR 0.08; p<0.0001), with an overall AUC of 0.91 (95% CI: 0.89-0.92). Amyloid status differences were statistically significant for Aβ42/40, GFAP, and NfL (p<0.05), with AUCs of 0.72 (95% CI: 0.70-0.75), 0.75 (95% CI: 0.72-0.77), and 0.55 (95% CI: 0.53-0.58), respectively. Although NfL demonstrated limited independent classification of amyloid status, it significantly contributed to classifying p-tau 217 borderline cases (Section 3.3).

**Figure 1.**
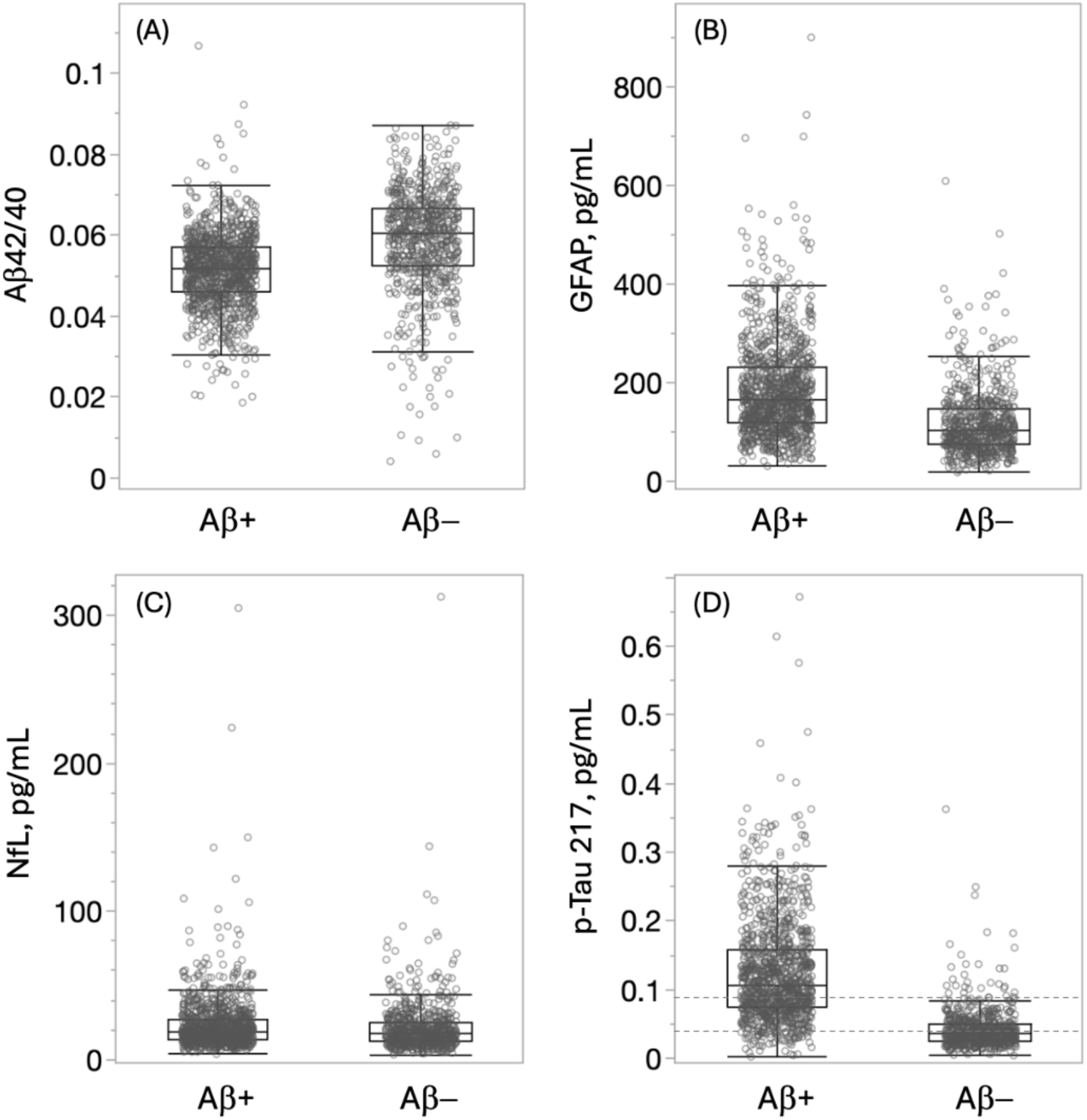
Biomarker levels (n-1,812) across training and validation cohorts, stratified by amyloid status. All biomarkers showed statistically significant differences between amyloid-positive and -negative groups. Dashed lines indicate the established intermediate p-tau 217 range (0.04-0.09 pg/mL). Continuous variables were used for all biomarkers in the algorithm, eliminating the need for discrete diagnostic thresholds for Ap42/40, GFAP, and NfL

### 3.3 Development of multi-analyte algorithm

We explored the potential benefit of combining p-Tau 217 detection with one or more of the N4PE biomarker assays, and found, as have others, that it is hard to improve upon p-Tau 217 detection for amyloid classification accuracy where p-Tau 217 is either low or high. While combining p-tau 217 with amyloid ratio or the full N4PE panel showed no statistically significant improvement in AUC in preliminary trials, we observed a stepwise increase in classification accuracy with more biomarkers specifically for cases within the intermediate range of previously optimized 2-cutoff thresholds for p-tau 217 [23]. To further explore multivariate modeling within this intermediate zone, we tested eight model types (Lasso regression, Bootstrap Forest, etc.) using various biomarker combinations, APOE status, sex, and age as inputs in an exploratory cohort of 735 symptomatic individuals (approximately 2/3 from the ADC and 1/3 from the Bio-Hermes cohort). Across model variations, AUCs remained statistically indistinguishable, averaging approximately 0.90, with Youden index-derived sensitivity and specificity converging around 85%. Given its simplicity and consistent performance, a nominal logistic regression model was selected for subsequent data fitting, with which we aimed to refine our understanding of how the additional biomarkers enhanced classification in the p-Tau 217 intermediate zone. Models were fit to predict amyloid status based on p-Tau 217 and Aβ42/40 ratio, and with all five biomarkers. Non-tau pathology markers (Aβ42/40 ratio, NfL, and GFAP) exhibited statistically significant, non-zero parameters in the models, with improved model metrics (R2, AIC, BIC) upon their inclusion. Notably, despite NfL’s limited standalone amyloid discrimination, it demonstrated the strongest statistical contribution to the all-biomarker multivariate model (p < 0.0001), surpassing Aβ42/40 ratio (p < 0.0048). Applying these 3- and 5-biomarker models to the exploratory cohort, we observed slight but non-significant increases in AUC (0.91 to 0.92) across models (Figure 2). The inset of Figure 2 highlights the midrange of the ROC curves where an incremental improvement in specificity with the addition of biomarkers suggested enhanced classification accuracy for cases within the p-tau 217 intermediate zone.

**Figure 2.**
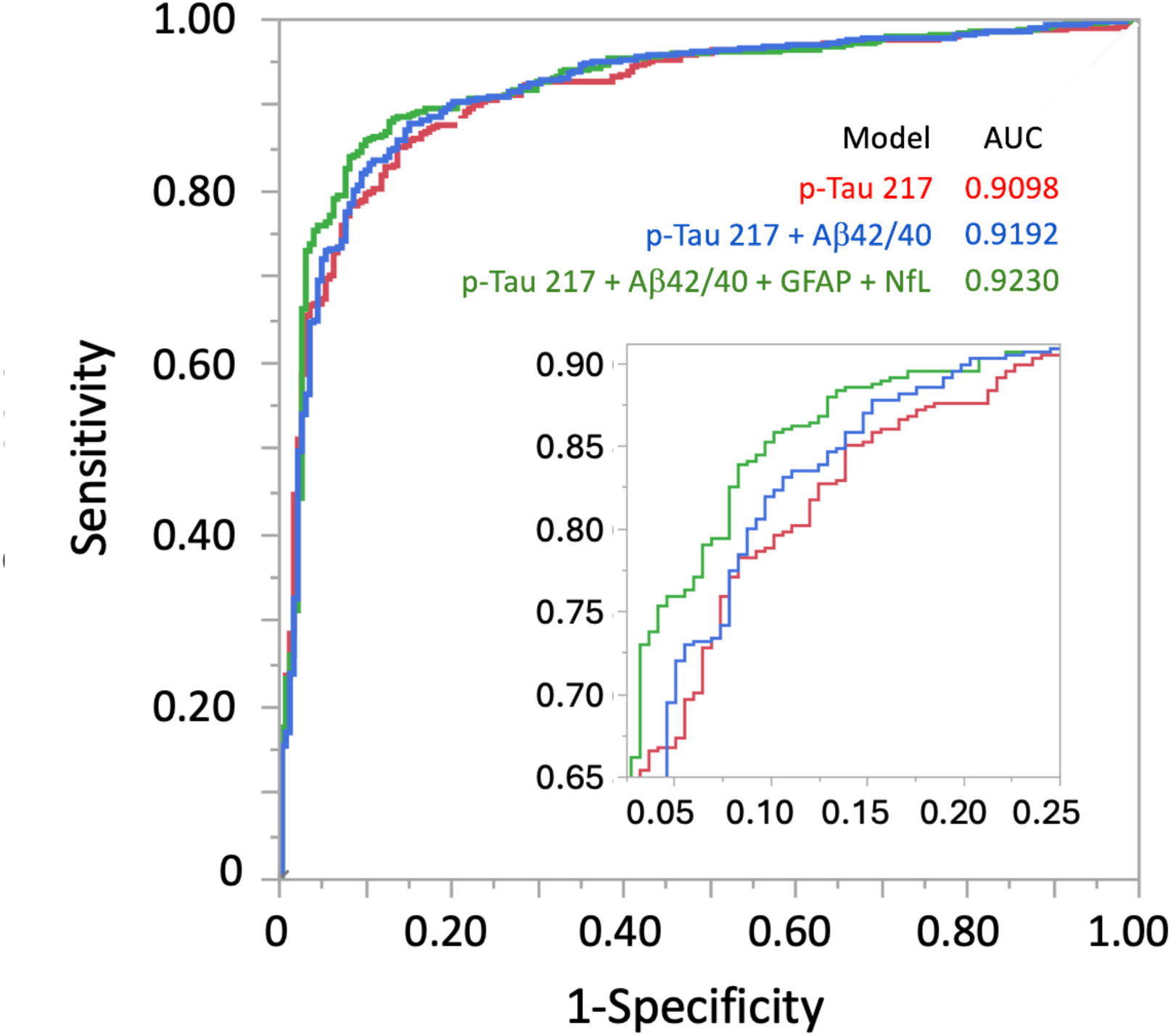
Receiver operating characteristic (ROC) curves comparing p-Tau 217 and multi-analyte models. AUC values were not statistically distinguishable. Inset: Zoomed-in view at 80% sensitivity, highlighting incremental increases in specificity that suggest improved classification accuracy within the p-Tau 217 intermediate range.

To better characterize the potential of the additional biomarkers to classify amyloid status in the p-Tau 217 intermediate zone, we derived thresholds from the best-fit sinh-arcsinh distributions of the multi-analyte score for amyloid-negative and -positive samples. These thresholds, targeting approximately 7% false negative and 7% false positive rates, yielded lower thresholds of 0.43 and 0.45, and upper thresholds of 0.74 and 0.70 for the 3- and 5-biomarker models, respectively. Compared to p-tau 217 alone, the intermediate zone decreased from 31.2% to 14.9% of samples in the exploratory sample set with the 3-biomarker model, and further to 10.5% with the 5-biomarker model (Table 2). This model’s stability and marker necessity were confirmed via forward stepwise selection, self-validating ensemble model, and multinomial regression, with all biomarkers demonstrating significant contributions (p < 0.0083 for GFAP, p < 0.0001 for others). Therefore, the 5-biomarker model was selected for subsequent validation.

**TABLE 2.** Comparison intermediate zones of p-Tau 217 and two multi-analyte models.

| **(A)** |  | pTau217 |  |  |  |  |  |  |
| --- | --- | --- | --- | --- | --- | --- | --- | --- |
|  |  | Low Risk |  | Intermediate |  | High Risk |  | All |
| Amyloid status |  | N | Row % | N | Row % | N | Row % | N |
| PET/CSF | Positive | 34 | 6.6% | 149 | 29.0% | 331 | 64.4% | 514 |
|  | Negative | 130 | 60.2% | 79 | 36.6% | 7 | 3.2% | 216 |
|  | All | 164 | 22.5% | 228 | 31.2% | 338 | 46.3% | 730 |
| **(B)** | | pTau217 + Aβ42/Aβ40 | | | | | | |
|  |  | Low Risk |  | Intermediate |  | High Risk |  | All |
| Amyloid status |  | N | Row % | N | Row % | N | Row % | N |
| PET/CSF | Positive | 42 | 8.2% | 69 | 13.4% | 403 | 78.4% | 514 |
|  | Negative | 158 | 73.1% | 40 | 18.5% | 18 | 8.3% | 216 |
|  | All | 200 | 27.4% | 109 | 14.9% | 421 | 57.7% | 730 |
| **(C)** | | pTau217 + Aβ42/Aβ40 + GFAP + NfL | | | | | | |
|  |  | Low Risk |  | Intermediate |  | High Risk |  | All |
| Amyloid status |  | N | Row % | N | Row % | N | Row % | N |
| PET/CSF | Positive | 46 | 8.9% | 41 | 8.0% | 427 | 83.1% | 514 |
|  | Negative | 162 | 75.0% | 36 | 16.7% | 18 | 8.3% | 216 |
|  | All | 208 | 28.5% | 77 | 10.5% | 445 | 61.0% | 730 |

Notes: Abbreviations: PET, positron emission tomography; CSF cerebrospinal fluid

To perform the test, samples are processed concurrently on the p-tau 217 and N4PE 4-plex assays. A multi-marker model score is calculated using a decision tree, initially stratifying samples based on p-tau 217 concentration. Samples with p-tau 217 concentrations below 0.04 pg/mL or above 0.09 pg/mL receive a logistic risk score derived solely from p-tau 217. Samples within the 0.04 to 0.09 pg/mL p-tau 217 range have their risk score determined by the logistic multi-analyte model, incorporating p-tau 217 and the N4PE biomarkers. This is represented by the equation:

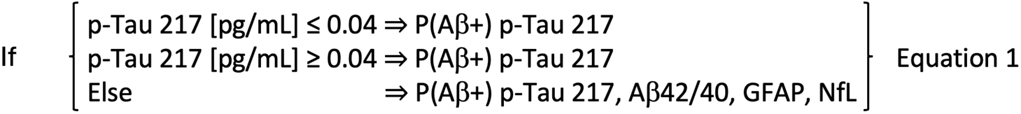

Where P(Aβ+) represents the probability of amyloid positivity, ranging from 0 to 1. Figure 3 illustrates the test’s workflow: identifying p-tau 217 intermediate zone cases (Figure 3A) and subsequently analyzing these cases with the multi-analyte algorithm. Although p-tau 217 concentrations are shown in the illustration, the test’s output for all samples is a risk score between 0 and 1. For user convenience, the risk score is multiplied by 100 and rounded to the nearest whole number.

**Figure 3.**
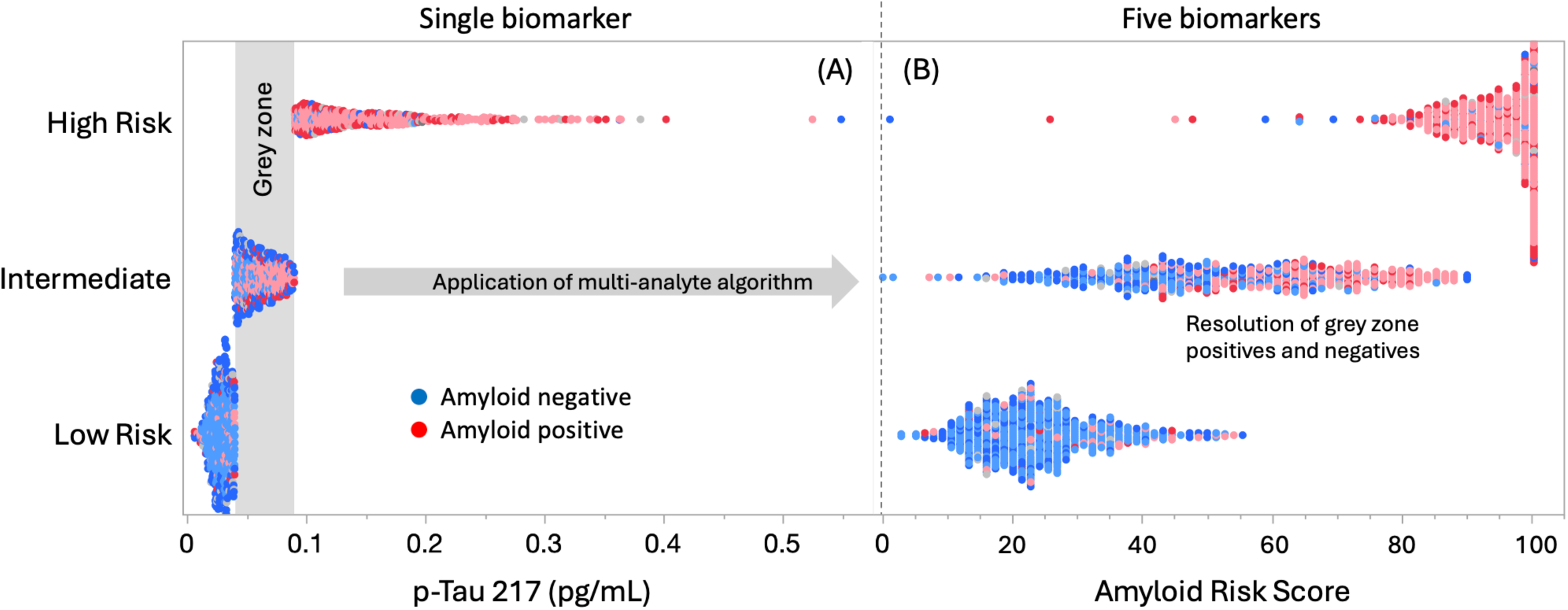
Multi-analyte interrogation of plasma samples for amyloid status. (A) p-Tau 217 classifies samples with low and high amyloid burden, identifying intermediate risk cases. (B) Multivariate analysis of all five biomarkers resolves approximately two-thirds of intermediate cases. All results are converted to an amyloid risk score for final interpretation.

### 3.4 Clinical performance validation

Clinical performance of the multi-analyte algorithmic test was assessed with the pre-defined thresholds described in Section 3.3. The validation datasets included the ADC cohort (n=274), Bio-Hermes validation sub-cohort (n=271), and the ADNI cohort (n=571), as detailed in Table 1. The overall prevalence of amyloid positivity was 55.4%. All 1,082 validation samples were tested in the Quanterix CLIA laboratory according to established laboratory procedures. In addition to the full sub cohort of ADNI samples, the individuals from this set having two or three longitudinal intervals from baseline in common (24 and 48 months) were analyzed separately at each timepoint as an assessment of the consistency of the test results across longitudinal samplings within individuals. Figure 4 depicts the results across the three validation cohorts. Figure 4A depicts an aggregate density plot, and risk score data are stratified by source and amyloid status in Figure 4B. The distributions of amyloid positive cases were qualitatively similar across the three cohorts, while there were more false positive results among the Bio-Hermes and ADNI cohorts than from the ADC cohort. It is unclear if the higher false positive rate may reflect lower sensitivity by visual PET (used in Bio-Hermes and ADNI) relative to CSF (used in ADC). Overall, 88% of the results were actionably outside of the intermediate grey zone.

**Figure 4.**
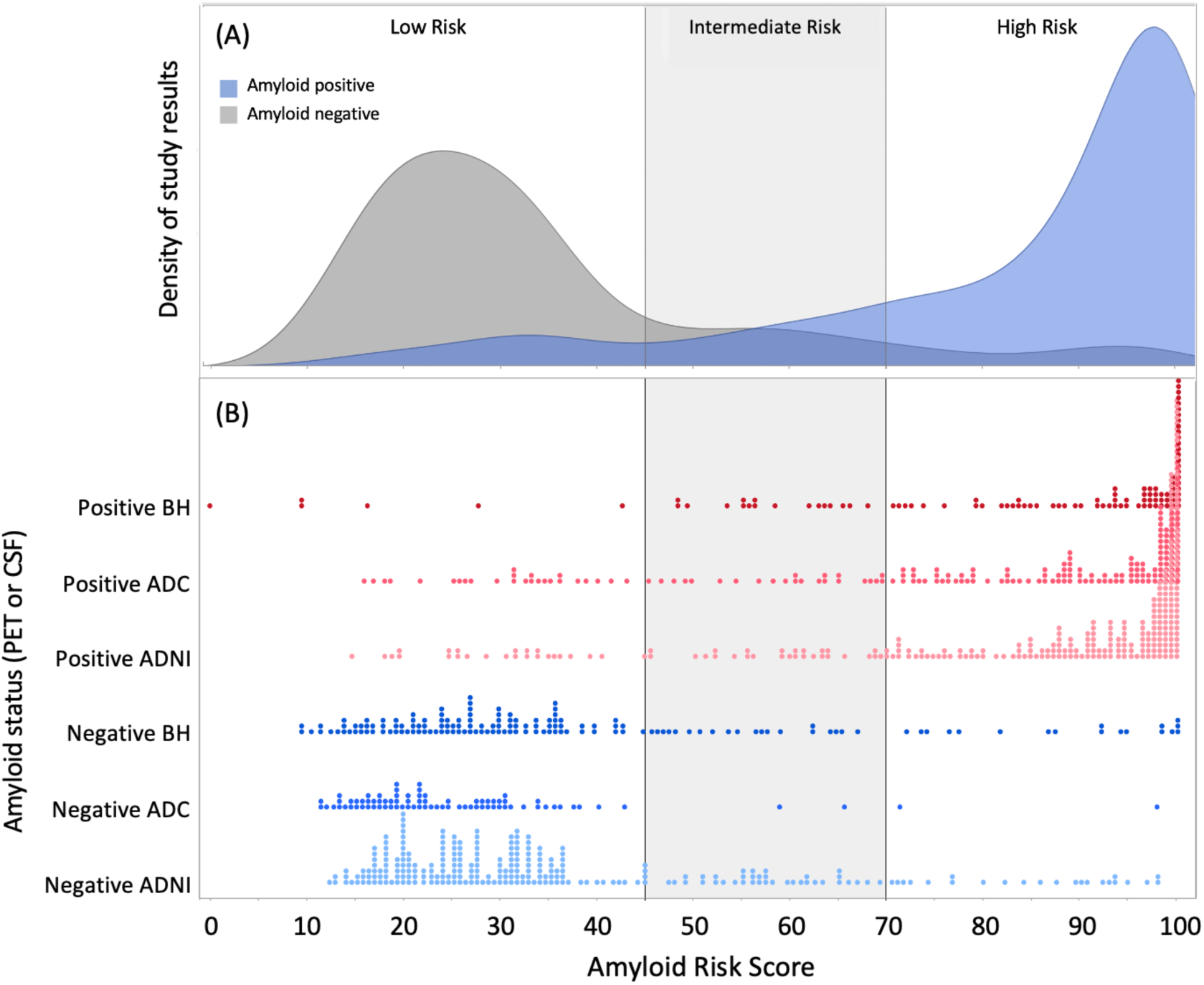
LucentAD Complete Amyloid Risk Score validation. (A) Distribution of test results across the full validation set. (B) Individual results by cohort and reference amyloid status, with the pre-established intermediate zone (45-70) highlighted in grey. Cohort abbreviations: BH, Bio-Hermes; ADC, Amsterdam Dementia Cohort; ADNI, Alzheimer’s Disease Neuroimaging Initiative.

Table 3 summarizes the clinical performance characteristics of the multi-analyte test. Overall, the combined validation statistics met the target accuracy of 90% and exhibited an intermediate range of 12%, well below the proposed maximum of 20% [3]. Importantly, the accuracy was statistically indistinguishable from the accuracy of p-Tau 217 alone as reported previously with a similar validation cohort, while the intermediate range was reduced from 30.9% [22]. The results demonstrate robust and reproducible performance across the diversity of the three cohorts and in relation to amyloid prevalence rates. This diversity included variations in race/ethnicity, age, geography, clinical settings, and reference methods. The test capability included maintaining positive predictive values (PPV) above 90% across amyloid prevalence rates ranging from 41% to 70%. Model-derived PPVs and negative predictive values (NPVs) across a broader range of amyloid prevalence rates, from 20% (cognitively normal older adults) to 65% (high-prevalence dementia population), are detailed in the supplemental materials. Tables 3A and 3B illustrate the effect of power to minimize the impact of variability in apparent performance metrics.

**TABLE 3.**
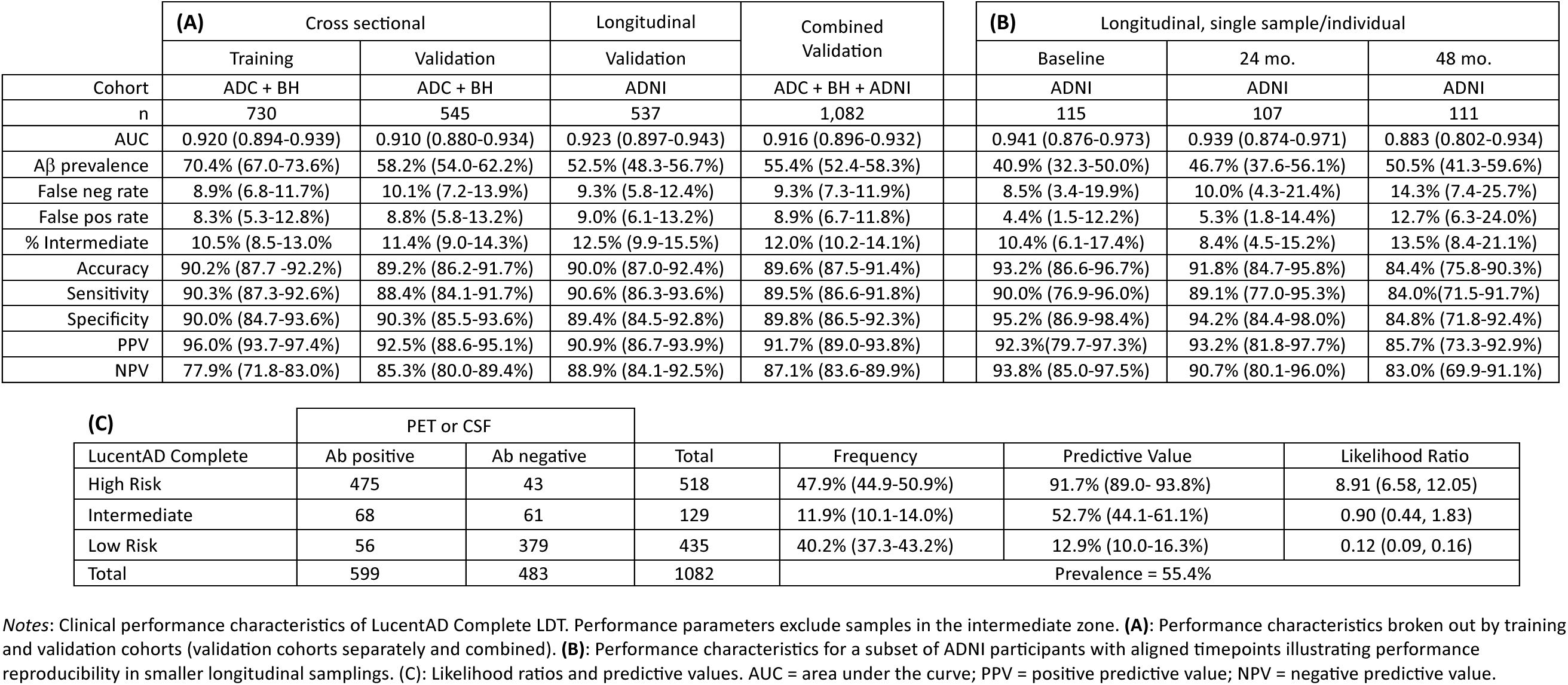
Clinical performance characteristics of LucentAD Complete.

|  | **(A)** Cross sectional |  | Longitudinal | Combined Validation | **(B)** Longitudinal, single sample/individual |  |  |
| --- | --- | --- | --- | --- | --- | --- | --- |
|  | Training | Validation | Validation |  | Baseline | 24 mo. | 48 mo. |
| Cohort | ADC + BH | ADC + BH | ADNI | ADC + BH + ADNI | ADNI | ADNI | ADNI |
| n | 730 | 545 | 537 | 1,082 | 115 | 107 | 111 |
| AUC | 0.920 (0.894-0.939) | 0.910 (0.880-0.934) | 0.923 (0.897-0.943) | 0.916 (0.896-0.932) | 0.941 (0.876-0.973) | 0.939 (0.874-0.971) | 0.883 (0.802-0.934) |
| Aβ prevalence | 70.4% (67.0-73.6%) | 58.2% (54.0-62.2%) | 52.5% (48.3-56.7%) | 55.4% (52.4-58.3%) | 40.9% (32.3-50.0%) | 46.7% (37.6-56.1%) | 50.5% (41.3-59.6%) |
| False neg rate | 8.9% (6.8-11.7%) | 10.1% (7.2-13.9%) | 9.3% (5.8-12.4%) | 9.3% (7.3-11.9%) | 8.5% (3.4-19.9%) | 10.0% (4.3-21.4%) | 14.3% (7.4-25.7%) |
| False pos rate | 8.3% (5.3-12.8%) | 8.8% (5.8-13.2%) | 9.0% (6.1-13.2%) | 8.9% (6.7-11.8%) | 4.4% (1.5-12.2%) | 5.3% (1.8-14.4%) | 12.7% (6.3-24.0%) |
| % Intermediate | 10.5% (8.5-13.0%) | 11.4% (9.0-14.3%) | 12.5% (9.9-15.5%) | 12.0% (10.2-14.1%) | 10.4% (6.1-17.4%) | 8.4% (4.5-15.2%) | 13.5% (8.4-21.1%) |
| Accuracy | 90.2% (87.7-92.2%) | 89.2% (86.2-91.7%) | 90.0% (87.0-92.4%) | 89.6% (87.5-91.4%) | 93.2% (86.6-96.7%) | 91.8% (84.7-95.8%) | 84.4% (75.8-90.3%) |
| Sensitivity | 90.3% (87.3-92.6%) | 88.4% (84.1-91.7%) | 90.6% (86.3-93.6%) | 89.5% (86.6-91.8%) | 90.0% (76.9-96.0%) | 89.1% (77.0-95.3%) | 84.0% (71.5-91.7%) |
| Specificity | 90.0% (84.7-93.6%) | 90.3% (85.5-93.6%) | 89.4% (84.5-92.8%) | 89.8% (86.5-92.3%) | 95.2% (86.9-98.4%) | 94.2% (84.4-98.0%) | 84.8% (71.8-92.4%) |
| PPV | 96.0% (93.7-97.4%) | 92.5% (88.6-95.1%) | 90.9% (86.7-93.9%) | 91.7% (89.0-93.8%) | 92.3% (79.7-97.3%) | 93.2% (81.8-97.7%) | 85.7% (73.3-92.9%) |
| NPV | 77.9% (71.8-83.0%) | 85.3% (80.0-89.4%) | 88.9% (84.1-92.5%) | 87.1% (83.6-89.9%) | 93.8% (85.0-97.5%) | 90.7% (80.1-96.0%) | 83.0% (69.9-91.1%) |

| **(C)** |  | PET or CSF |  | Total | Frequency | Predictive Value | Likelihood Ratio |
| --- | --- | --- | --- | --- | --- | --- | --- |
| LucentAD Complete |  | Ab positive | Ab negative |  |  |  |  |
| High Risk |  | 475 | 43 | 518 | 47.9% (44.9-50.9%) | 91.7% (89.0- 93.8%) | 8.91 (6.58, 12.05) |
| Intermediate |  | 68 | 61 | 129 | 11.9% (10.1-14.0%) | 52.7% (44.1-61.1%) | 0.90 (0.44, 1.83) |
| Low Risk |  | 56 | 379 | 435 | 40.2% (37.3-43.2%) | 12.9% (10.0-16.3%) | 0.12 (0.09, 0.16) |
| Total |  | 599 | 483 | 1082 | Prevalence = 55.4% |  |  |

*Notes:* Clinical performance characteristics of LucentAD Complete LDT. Performance parameters exclude samples in the intermediate zone. **(A):** Performance characteristics broken out by training and validation cohorts (validation cohorts separately and combined). **(B):** Performance characteristics for a subset of ADNI participants with aligned timepoints illustrating performance reproducibility in smaller longitudinal samplings. **(C):** Likelihood ratios and predictive values. AUC = area under the curve; PPV = positive predictive value; NPV = negative predictive value.

Table 3C presents likelihood ratios: a high likelihood ratio for high-risk test results calls (lower 95% CI > 6) indicating a strong positive signal for amyloid-positive cases, a likelihood ratio close to 1 for the intermediate zone suggesting no significant difference in positive and negative test results, and a low likelihood ratio of 0.12 for low-risk test results calls indicating a much higher likelihood of negative calls in amyloid-negative individuals than false-positive calls.

### 3.5 Performance in subgroups

A detailed analysis of the LucentAD Complete test’s performance in patients with dementia due to other etiologies (VaD, FTD, DLB) and co-pathology is provided in the supplemental materials. These subgroups showed amyloid positivity rates of 36.0% (VaD), 19.8% (FTD), and 47.6% (DLB) by CSF, consistent with expected prevalence ranges for these dementia syndromes [26]. Amyloid detection accuracies for the LucentAD Complete test were 90.6% for VaD, 87.3% for FTD, and 76.9% for DLB. While detection accuracy for AD pathology in VaD and FTD was statistically consistent to the combined validation cohorts, the accuracy for DLB was significantly lower. In addition, the percentage of cases in the intermediate zone (25%) in DLB patients was statistically higher than for the other two dementia types and the overall validation estimate (12%). Next, inclusion of these co-pathology cases into the validation sample set at expected proportions for a memory clinic population [26–31] did not significantly alter the test’s amyloid classification performance (see supplemental materials). Except for DLB, accuracy of amyloid detection across demographic subgroups ranged from 87% to 95% with the percent of intermediate results ranging from 3.5% to 14% (Figure 5). Although the accuracy estimate for non-White participants (95%) was numerically higher than for White participants, this difference was not statistically significant. Similarly, age, sex, and APOE4 carrier status did not significantly impact test accuracy (Figure 5).

**Figure 5.**
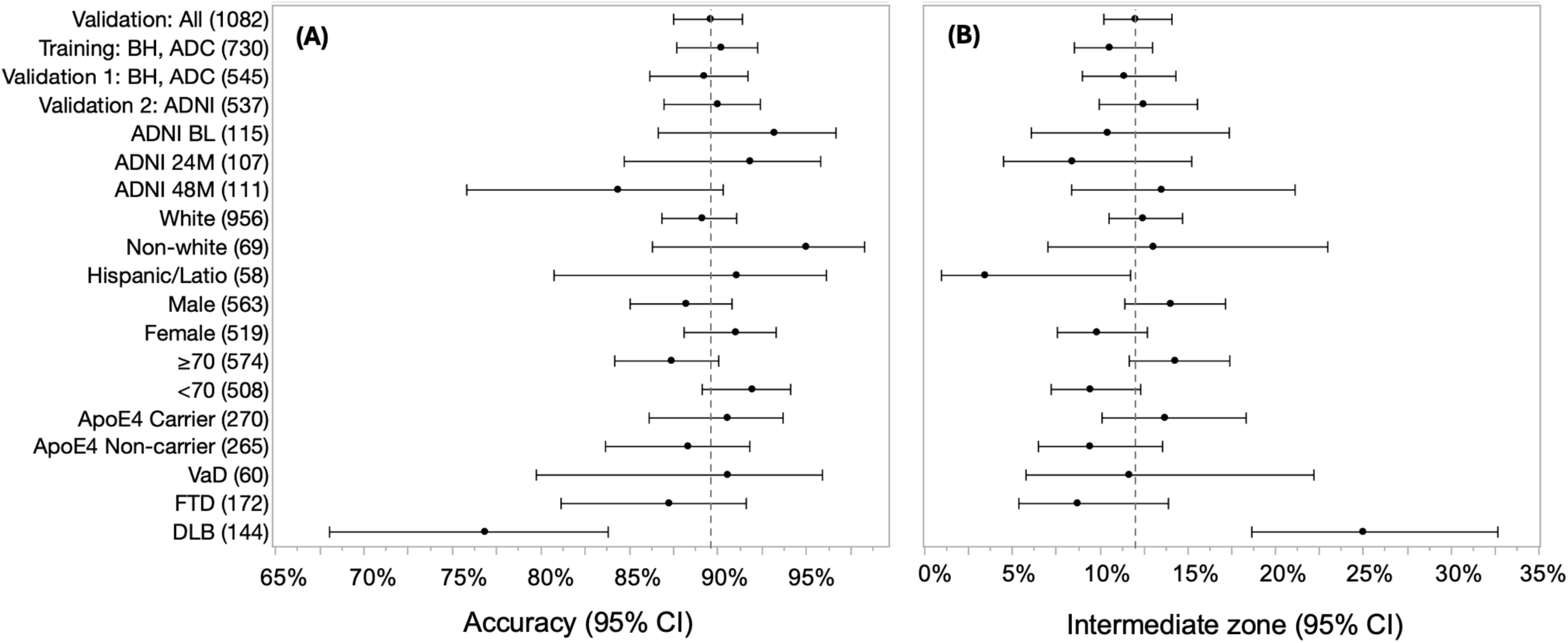
LucentAD Complete accuracy (A) and intermediate zone percentages (B) across datasets and subgroups. Point estimates and 95% confidence intervals are shown for training, validation, and subgroup analyses. Vertical lines represent the full validation set (n=l,082). Sample sizes are in parentheses. Non-white subgroup: Black, Asian, and Native American participants. Cohort/subgroup abbreviations: Validation 1, first validation cohort; Validation 2, second validation cohort; BH, Bio­Hermes; ADC, Amsterdam Dementia Cohort; ADNI, Alzheimer’s Disease Neuroimaging Initiative; VaD, vascular dementia; FTD, frontotemporal dementia; DLB, dementia with Lewy bodies; BL, baseline; 24M, 24 months; 48M, 48 months.

## 4 DISCUSSION

This multi-cohort study demonstrates the clinical validity of the LucentAD Complete test, a novel blood-based assay for amyloid pathology detection. Our findings build upon the established value of p-Tau 217 as a highly accurate single biomarker but also address the persistent challenge of classifying individuals within the p-Tau 217 intermediate zone. This multi-analyte approach demonstrated an AUC of 0.92 and 90% accuracy, while reducing the intermediate zone approximately 3-fold to 12%. This improvement enhances diagnostic confidence and potentially reduces the need for more invasive and costly procedures like CSF analysis or PET scans. While p-Tau 217 alone offers excellent discrimination for many individuals, the presence of an indeterminate zone necessitates further investigation, a challenge addressed by this novel assay.

Our key finding is that the addition of four readily measurable plasma biomarkers (Aβ42/Aβ40, GFAP, and NfL) significantly enhances amyloid classification specifically within this critical intermediate zone. This improvement can be rationalized by the multifactorial nature of Alzheimer’s disease, which extends beyond amyloid plaque deposition and p-Tau production [32]. Variable levels of inflammation and neuronal damage across individuals suggest that a broader assessment using biomarkers representing these diverse pathological processes improves the identification of patients exhibiting multiple disease-related biomarkers [33]. Importantly, this enhancement is achieved without compromising the high accuracy of p-Tau 217 in clearly classified amyloid-positive or -negative samples. This targeted approach, utilizing a multi-analyte algorithm solely within the p-Tau 217 intermediate range, optimizes the LucentAD Complete test for diagnostic power while minimizing complexity. The observed 3-fold reduction in the intermediate zone (31.2 to 10.5% in the training set) translates to a greater proportion of patients receiving clear and actionable results, thereby improving clinical workflow and potentially accelerating access to appropriate interventions.

The multi-analyte algorithm and established cut-offs demonstrated robust performance across three independent cohorts, comprising diverse demographics, clinical characteristics, and amyloid positivity prevalence rates. This heterogeneity, including variations in age, geography, race/ethnicity, and clinical presentation, strengthens the generalizability of our findings and suggests that LucentAD Complete can be reliably applied across a broad spectrum of clinical settings. Notably, the test maintained a positive predictive value exceeding 90% across amyloid prevalence rates ranging from 41% to 70%, a critical attribute for clinical utility. The inclusion of the Bio-Hermes cohort, designed to enrich for racial and ethnic diversity, is an important step towards ensuring equitable access to advanced diagnostic tools. Further studies will be needed to explore the test’s performance across even more diverse populations. The longitudinal analysis within the ADNI cohort provides preliminary evidence for the test’s consistency over time, an important consideration for potentially monitoring disease progression.

Several limitations should be acknowledged. First, the LucentAD Complete test exhibited reduced accuracy in detection AD pathology with the DLB patient group, with diminished sensitivity and a higher proportion of cases falling within the intermediate zone. This suggests that the amyloid signal in DLB patients may be more challenging to detect in plasma, potentially resulting in a high positive predictive value but a lower negative predictive value in this population. Therefore, while a positive result strongly indicates amyloid co-pathology in DLB, a negative result may not reliably rule it out. This finding should be considered when interpreting test results in patients with suspected DLB. Additionally, the use of different reference standards (CSF biomarkers in ADC, PET scans in Bio-Hermes and ADNI) could contribute to some of the observed differences in test performance across cohorts, particularly the apparent higher false positive rate in the PET-based cohorts. While visual assessment of PET scans is the current standard, it is possible that quantitative PET analysis or other advanced imaging techniques could provide more granular and consistent measures of amyloid burden. Quantitative PET demonstrates higher sensitivity and consistency than visual PET [34], and CSF biomarkers exhibit superior clinical performance compared to amyloid PET, with a 22% discordance rate that was predominantly CSF+/PET- [35]. Furthermore, this study included only cognitively impaired individuals (MCI, mild AD), and further work is needed to evaluate the test in asymptomatic populations, especially with emerging pre-clinical AD treatments [36].

While this study focused on evaluating the diagnostic performance of LucentAD Complete for amyloid pathology, future research should investigate its potential utility in other clinical contexts. Investigating its potential in predicting conversion from MCI to AD or in identifying individuals most likely to benefit from specific therapies would be valuable. Additionally, clinical utility and cost-effectiveness analyses will be essential to fully understand the impact of LucentAD Complete on clinical practice and healthcare resource utilization. These studies are underway, including the multi-site Accurate Diagnosis blood-based biomarker implementation study sponsored by the Davos Alzheimer’s Collaborative [37]. Finally, the availability of results from all four biomarkers, particularly NfL, opens the possibility for extending the test’s application to help guide differential diagnosis. For instance, patients exhibiting low amyloid risk, but elevated NfL levels may suggest a diagnostic path toward non-AD neurodegenerative pathologies, such as FTD [17, 38] or VaD.

In summary, LucentAD Complete marks a significant advancement in blood-based amyloid detection. By integrating p-Tau 217 with a panel of complementary biomarkers and employing a targeted algorithmic approach, this test offers high accuracy and a substantially reduced intermediate zone, facilitating more efficient and accessible Alzheimer’s disease diagnosis. The consistent and robust performance observed across diverse cohorts underscores its potential clinical utility as a valuable tool for aiding in the evaluation of individuals with suspected Alzheimer’s disease.

## Supporting information

Supplemental materials.doc

## Data Availability

All data produced in the present study are available upon reasonable request to the authors

## Funding Statement

Charlotte E. Teunissen is a recipient of TAP-dementia (www.tap-dementia.nl), receiving funding from ZonMw (#10510032120003) in the context of Onderzoeks programma Dementie, part of the Dutch National Dementia Strategy and ABOARD, which is a public-private partnership receiving funding from ZonMW (#73305095007) and Health∼Holland, Topsector Life Sciences & Health (PPP-allowance; #LSHM20106). Alzheimer NederlandInge. Prof. Teunissen further received grants of the European Commission (Marie Curie International Training Network, grant agreement No 860197 (MIRIADE), Innovative Medicines Initiatives 3TR (Horizon 2020, grant no 831434) EPND (IMI 2 Joint Undertaking (JU), grant No. 101034344), and JPND (bPRIDE), National MS Society (Progressive MS Alliance), Alzheimer Drug Discovery Foundation, Alzheimer Association, Health Holland, the Dutch Research Council (ZonMW), the Selfridges Group Foundation, and Alzheimer Netherlands. M.W. Verberk is supported by grants of the Alzheimer’s Association, Health∼Holland and Amsterdam UMC.

## Conflict of Interest

Charlotte E. Teunissen performed contract research for Acumen, ADx Neurosciences, AC-Immune, Alamar, Aribio, Axon Neurosciences, Beckman-Coulter, BioConnect, Bioorchestra, Brainstorm Therapeutics, Celgene, Cognition Therapeutics, EIP Pharma, Eisai, Eli Lilly, Fujirebio, Grifols, Instant Nano Biosensors, Merck, Novo Nordisk, Olink, PeopleBio, Quanterix, Roche, Toyama, Vivoryon. Charlotte E. Teunissen is editor in chief of Alzheimer Research and Therapy, and serves on editorial boards of Medidact Neurologie/Springer, and Neurology: Neuroimmunology & Neuroinflammation. Inge Verberk received a speaker honorarium from Quanterix, which was paid directly to her institution. David Wilson, Meenakshi Khare, Michele Wolfe, Patrick Sheehy, Ann-Jeanette Vasko, and Mike Miller are employees of Quanterix. Karen Copeland and Lyndal Hesterberg are contractors of Quanterix.

## Ethics Statement

All studies were properly consented and approved by applicable ethics committees. The studies were conducted in accordance with local legislation and institutional requirements.

## Notes

### Author Declarations

All Amsterdam Dementia Cohort participants provided written informed consent to use medical data and biomaterials for research purposes. The study is approved by the medical ethical committee of the VU University Medical Center (Ref: 2016.061, Ref: 2017.315), and is conducted in accordance with the Helsinki Declaration of 1975. The BioHermes study and informed consent was reviewed and approved by Advarra, 6100 Merriweather Dr., Suite 600 Columbia, MD 21044. The Alzheimer's Disease Neuroimaging Initiative (ADNI) study procedures were approved by the institutional review boards (IRBs) of all participating centers, and written informed consent was obtained from all participants or their authorized representatives.

