## Supplemental materials.doc for "Clinical Performance of LucentAD Complete: A Multi-Analyte Algorithmic Blood Test for Detecting Brain Amyloid with a Scalable, Multiplexed Digital Immunoassay"

**Table of Contents**

.

A. Combined validation cohort demographics stratified by amyloid status ……………………….	2
B. Analysis of VaD, FTD, DLB dementia samples…………….......................................................3
C. Evaluation of precision of amyloid risk score ………….........................................................	6
D. Model-derived PPV, NPVs across amyloid prevalence rates…………………………………………..	8

**A.** Table S1: Combined validation cohort demographics stratified by amyloid status

|  | Aβ+ (N = 599) | Aβ− (N = 483) | All (N = 1082) |
| --- | --- | --- | --- |
| **Age** |  |  |  |
| Mean (SD) | 70.6 (6.8) | 69.0 (7.7) | 69.9 (7.8) |
| Range | 45-86 | 44-86 | 44-86 |
| **Sex** |  |  |  |
| F | 295 (49.2%) | 224 (46.4%) | 519 (48.0%) |
| M | 304 (50.8%) | 259 (53.6%) | 563 (52.0%) |
| **Race** |  |  |  |
| Native American or Alaskan | 0 (0.0%) | 3 (0.6%) | 3 (0.3%) |
| Asian | 7 (3.0%) | 8 (1.7%) | 15 (1.3%) |
| Black or African American | 16 (2.7%) | 31 (6.4%) | 47 (4.3%) |
| More than one race | 3 (0.5%) | 1 (0.2%) | 4 (0.4%) |
| Unknown | 0 (1.5%) | 4 (0.8%) | 4 (0.4%) |
| White | 573 (95.7%) | 436 (90.3%) | 1,009 (93.3%) |
| **Ethnicity** |  |  |  |
| Hispanic or Latino | 32 (5.3%) | 26 (5.4%) | 58 (5.4%) |
| Not Hispanic or Latino | 566 (94.5%) | 457 (94.6%) | 1,023 (94.5%) |
| Not Reported | 1 (0.2%) | 0 (0.0%) | 1 (0.1%) |
| **MMSE** |  |  |  |
| Mean (SD) | 24.4 (4.8) | 27.2 (2.6) | 25.6 (4.1) |
| Range | 2-30 | 16-30 | 2-30 |
| Missing | 50 (0.0%) | 72 (1.5%) | 122 (11.3%) |
| **Diagnostic Group** |  |  |  |
| AD | 177 (29.5%) | 12 (2.5%) | 189 (17.5%) |
| Mild AD | 74 (12.4%) | 48 (9.9%) | 122 (11.3%) |
| MCI | 348 (58.1%) | 423 (87.6%) | 771 (71.3%) |
| **ApoE** |  |  |  |
| ε2ε3 | 13 (2.2%) | 68 (14.1%) | 81 (7.6%) |
| ε2ε4 | 16 (2.7%) | 4 (0.8%) | 20 (1.9%) |
| ε3ε3 | 205 (34.2%) | 319 (66.0%) | 524 (48.9%) |
| ε3ε4 | 276 (46.1%) | 75 (15.5%) | 351 (32.7%) |
| ε4ε4 | 86 (14.4%) | 10 (2.1%) | 96 (9.0%) |
| ε4 carriers | 378 (63.1%) | 89 (18.4%) | 467 (43.6%) |
| Missing | 3 (0.5%) | 7 (1.4%) | 10 (0.9%) |
| **Amyloid Status** |  |  |  |
| CSF | 188 (31.4%) | 75 (15.5%) | 263 (24.3%) |
| PET | 411 (68.6%) | 408 (84.5%) | 819 (75.7%) |
| Amyloid Risk Score (SD) | 83.4 (22.3) | 35.0 (20.8) | 61.8 (32.4) |
| Amyloid Risk Score, range | 0.00-1.00 | 9.5-1.00 | 0-1.00 |

*Notes*: Validation cohorts. Abbreviations: BH, Bio-Hermes; ADC, Amsterdam Dementia Cohort; ADNI, Alzheimer’s Disease Neuroimaging Initiative; MCI, mild cognitive impairment; PET, positron emission tomography.

**B.** Analysis of VaD, FTD, DLB dementia samples

Sub cohorts of cases diagnosed with VaD (n=60), FTD (n=172), and DLB (n=144) were tested in the LucentAD Complete assay. A proportion of these samples were also amyloid positive, and the accuracy of the test for detection of amyloid in these mixed pathology cases was characterized. Demographic and clinical characteristics of these samples are summarized in Table S2.

**Table S2: Characteristics of VaD, FTD, and DLB samples from the ADC**

|  | **VaD** | **FTD** | **DLB** |
| --- | --- | --- | --- |
| n | 60 | 172 | 144 |
| Age (mean, SD) | 67.5 (7.4) | 64.3 (6.7) | 69.0 (6.5) |
| Sex (female) | 41 (82.0) | 47% | 19% |
| APOE carrier = yes (%) | NA | 31% | 57% |
| MMSE (mean, SD) | NA | 23.5 (4.9) | 22.3 (4.7) |
| CSF Abeta42 (mean, SD) | 798.7 (214.1) | 1005.9 (333.5) | 807.7 (266.6) |
| CSF p-Tau 181 (mean, SD) | 43.5 (21.6) | 48.8 (20.6) | 53.3 (23.6) |
| CSF Tau (mean, SD) | 330.4 (200.8) | 402.7 (193.7) | 390.1 (243.7) |
| Amyloid positive by CSF (%) | 30.0% | 19.2% | 48.6% |

LucentAD Complete results compared with CSF amyloid status for the VaD, FTD, and DLB cases are summarized in Tables S3-S5 below.

Table S3: 2 x 3 Table for VaD cases

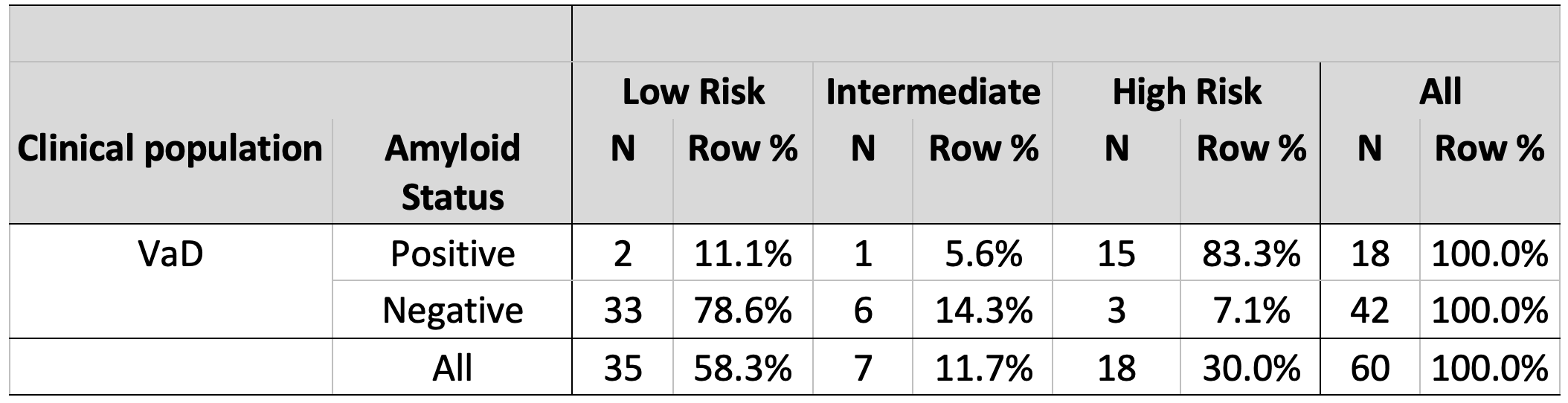

Table S4: 2 x 3 Table for FTD cases

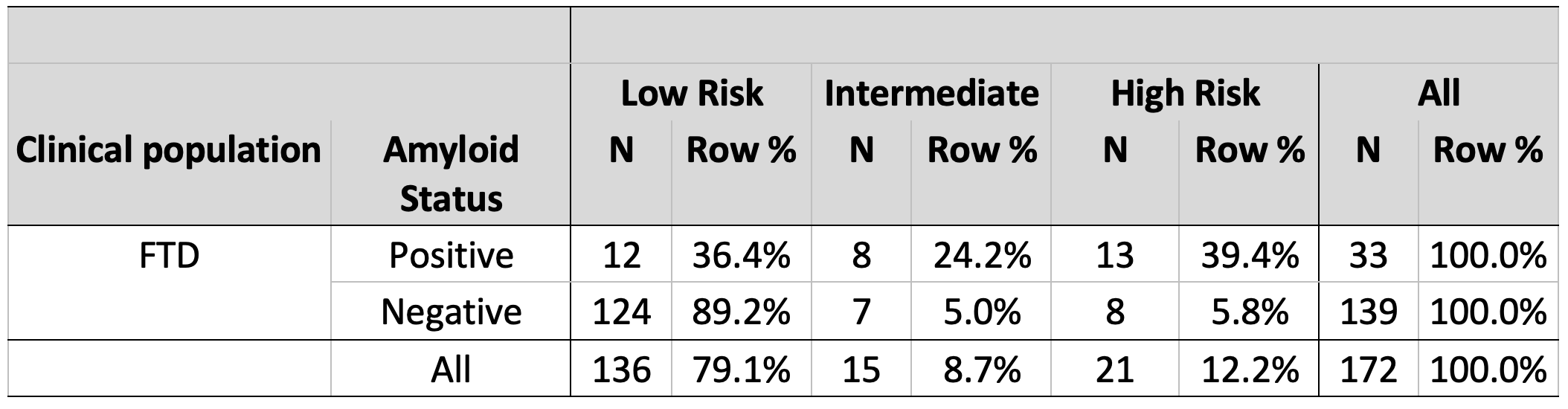

Table S5: 2 x 3 Table for DLB cases

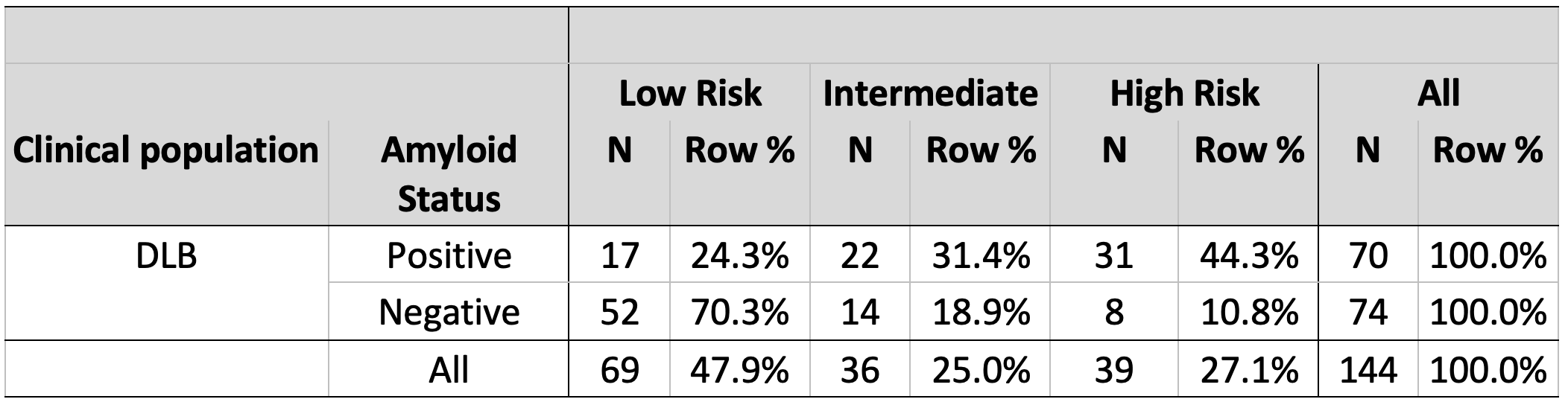

Clinical performance metrics obtained for the VaD, FTD, and DLB cases are summarized in Table S6:

Table S6: Performance metrics with 95% CI for DLB, FTD, and VaD cases

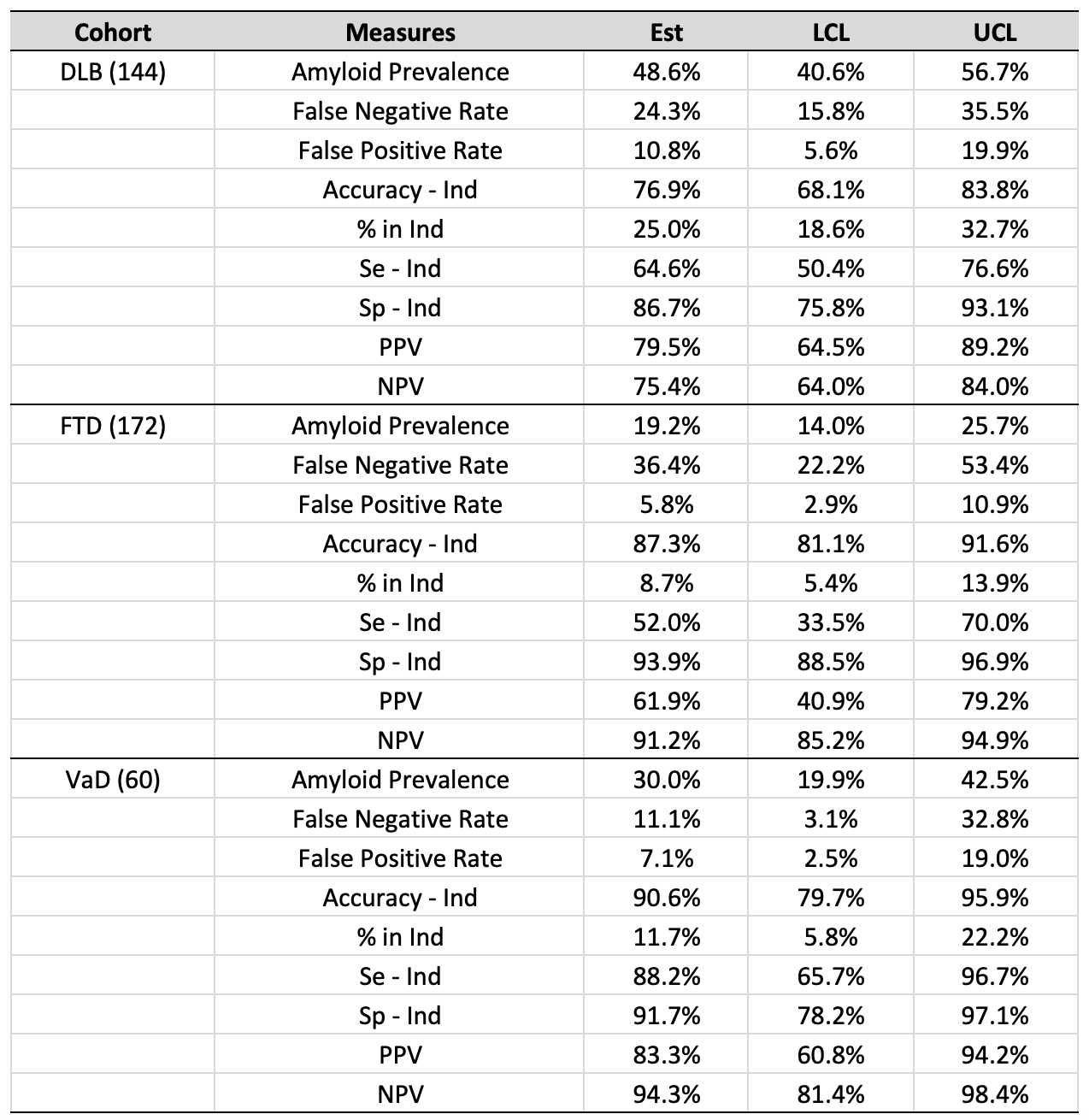

The data indicate that brain amyloid pathology detection accuracies in FTD and VaD cases are statistically consistent with the accuracy obtained with the combined validation cohort, however, the accuracy of amyloid detection in DLB cases (76.9%) was statistically lower. The proportion of FTD and VaD cases falling in the intermediate zone was also statistically consistent with the combined validation cohort results, while the proportion of DLB cases is statistically higher. These results suggest the amyloid signal in plasma in DLB cases is weaker than in other non-AD dementia categories.

The impact of inclusion of these non-AD and co-pathology cases into the combined validation cohort (n=1,082) was assessed. Two different incidence levels were examined: “typical” percentages as reported in memory clinics, and “high” levels as might be encountered with under-diagnoses and among a younger population with high percentages of FTD. Table S7 summarizes the percentages that were included:

**Table S7: Percentage of VaD, FTD, and DLB cases included in validation cohort**

|  | Non-AD dementia samples included in validation cohort | Validation cohort n with non-AD samples added | % of each non-AD dementia diagnostic group in validation cohort |
| --- | --- | --- | --- |
| Low | 60 VaD/60 FTD/60 DLB | 1,262 | 4.8% VaD/4.8% FTD^27-30^/4.8%^26^ DLB |
| High | 60 VaD/120 FTD/120 DLB | 1,382 | 4.3% VaD/8.7% FTD^31^/8.7% DLB |

Only a maximum of 4.8% of VaD cases could be included due to a limitation on the number of available samples (60). However, because high accuracy for amyloid detection in VaD cases was observed (90.6%), inclusion of additional VaD samples is not expected to alter the overall diagnostic performance of the test.

The effect of the addition of up to 300 non-AD dementia cases is depicted in Figure S8 below.

**Figure S8: Effect of the addition of VaD, FTD, & DLB cases on clinical performance metrics**

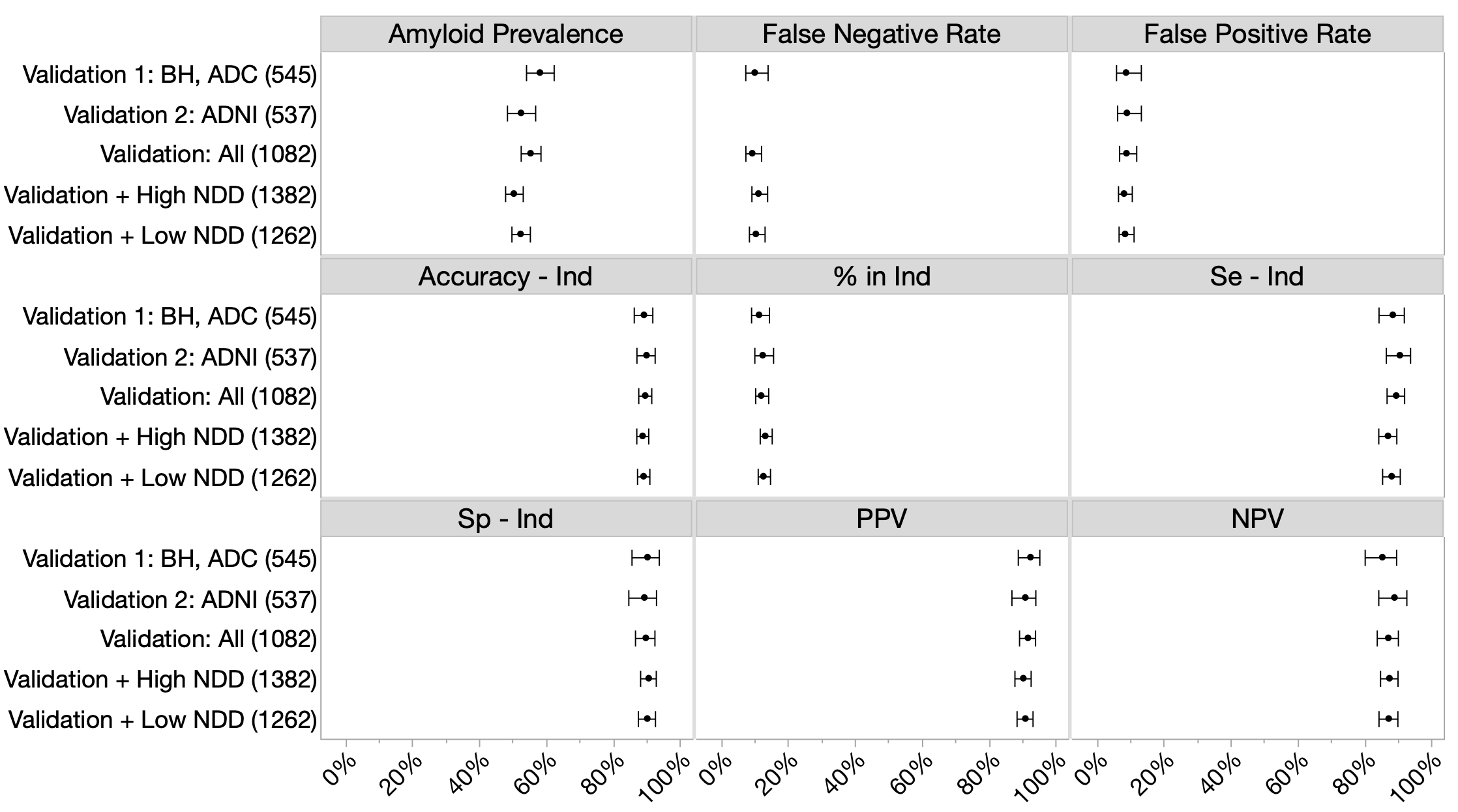

*Notes*: Clinical performance parameters for sub cohorts (Validation 1, Validation 2) and the combined validation cohort (“All”) are depicted. Abbreviations: NDD=neurodegeneration samples (VaD, FTD, DLB), BH=Bio-Hermes, ADC=Amsterdam Dementia Cohort, ADNI=Alzheimer’s Disease Neuroimaging Initiative.

Despite the lower accuracy for amyloid detection in DLB cases, there was no significant difference in the clinical performance of LucentAD Complete in classifying amyloid status with up to 22% (300/1,382) of non-AD dementia cases (Table S9).

**Table S9: Effect of the addition low and high levels of non-AD dementia samples on clinical performance metrics**

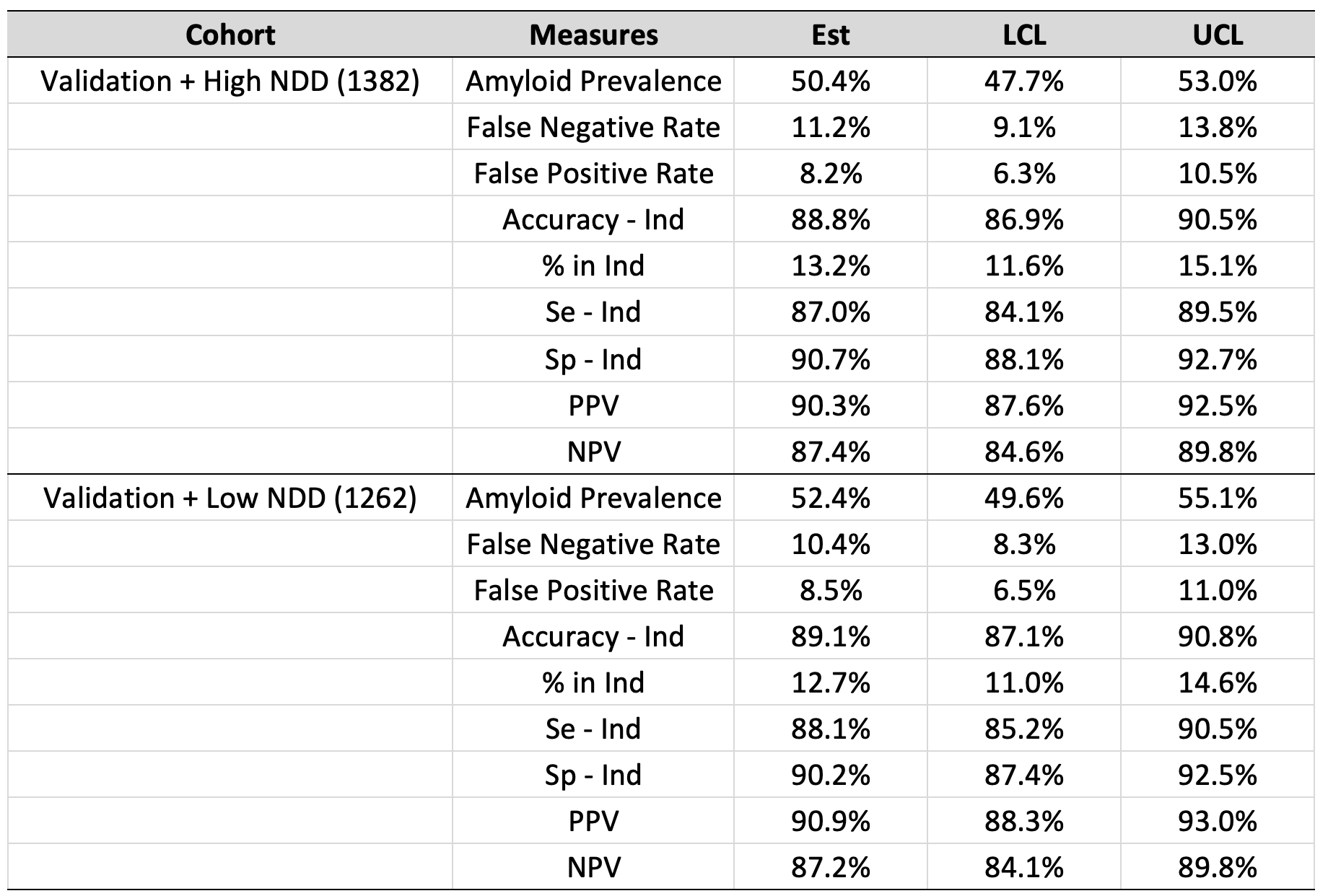

*Notes*: NDD=neurodegeneration samples (VaD, FTD, DLB).

**C.** Evaluation of Precision of LucentAD Complete Amyloid Risk Score

The precision of the LucentAD Complete logistic risk score relies on the precision of each constituent assay. LucentAD Complete measures five analytes. Notably, four of these (Aβ40, Aβ42, GFAP, and NfL) are measured within a single multiplexed immunoassay, minimizing the additive variability that would arise from separate immunoassays. This consolidation enhances the overall precision of the algorithm. To evaluate the impact of logistic risk score imprecision on amyloid classification accuracy, we performed a modeling study. This involved: 1) estimating biomarker imprecision through a multi-day precision study (**Figure S10**); 2) defining a normal error distribution for each biomarker using the precision study's variance estimates; 3) randomly adding or subtracting error from each biomarker; 4) calculating logistic multi-marker scores using the simulated biomarker values with added error; 5) conducting 10 simulations for each of 1,305 samples, mimicking 10 runs per sample in a laboratory setting (**Figure S11**); and 6) assessing the resulting risk score imprecision and its effect on amyloid classification accuracy.

**Figure S10: CV profiles of the five biomarkers**

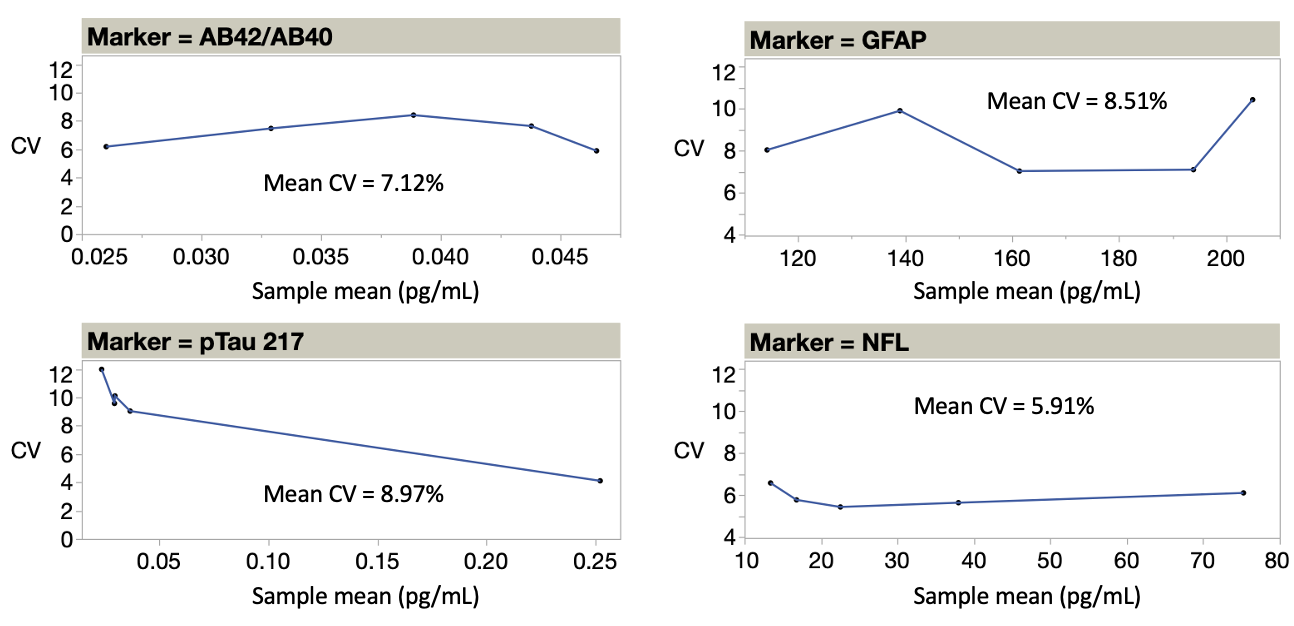

**Fig S10**. Precision profiles of each of the five biomarkers (two biomarkers represented the amyloid ratio) tested at five different levels across five days, two runs/day.

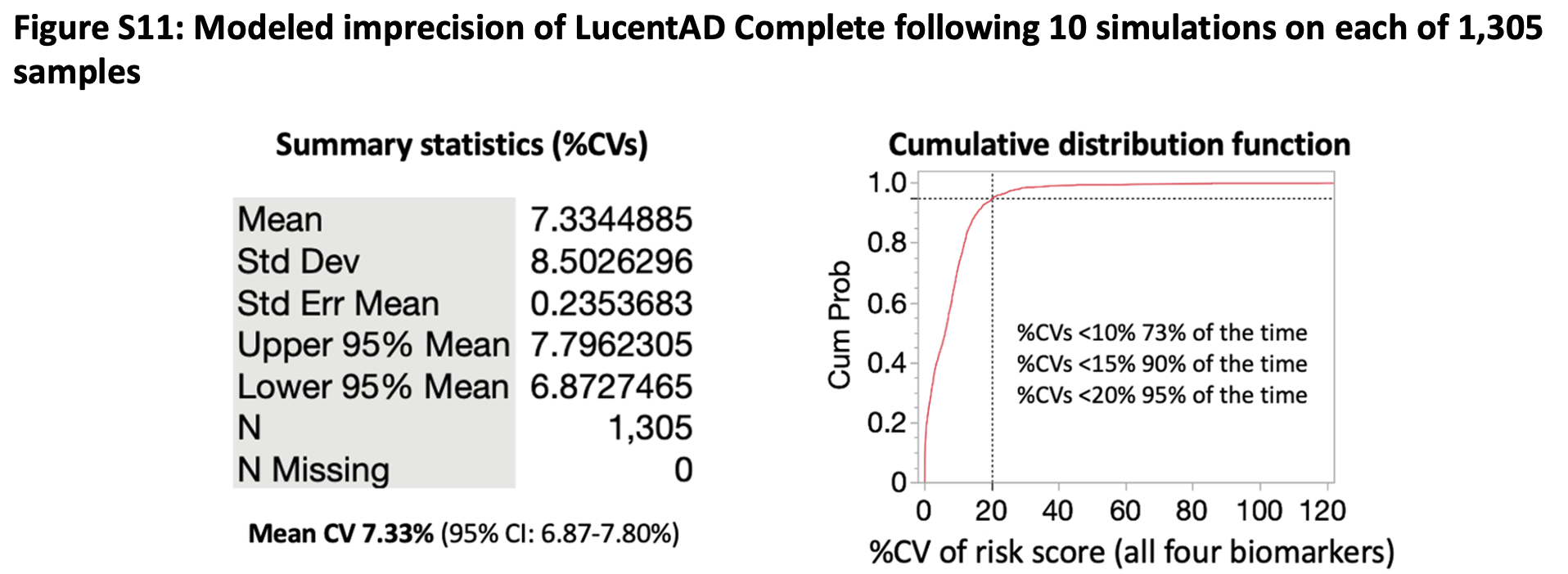

**Fig S11**. Modeled imprecision of LucentAD Complete risk score across 13,050 error simulations based on actual variances of each biomarker.

As shown by **Figure S11**, the overall mean imprecision (%CV) of the test across all simulations was **7.33%** (95% CI: 6.87-7.80%). At this level of precision relative to clinical cutoffs, the potential low to high or high to low amyloid risk misclassification was estimated to be 1%.

D. Model-derived PPV, NPVs across amyloid prevalence rates

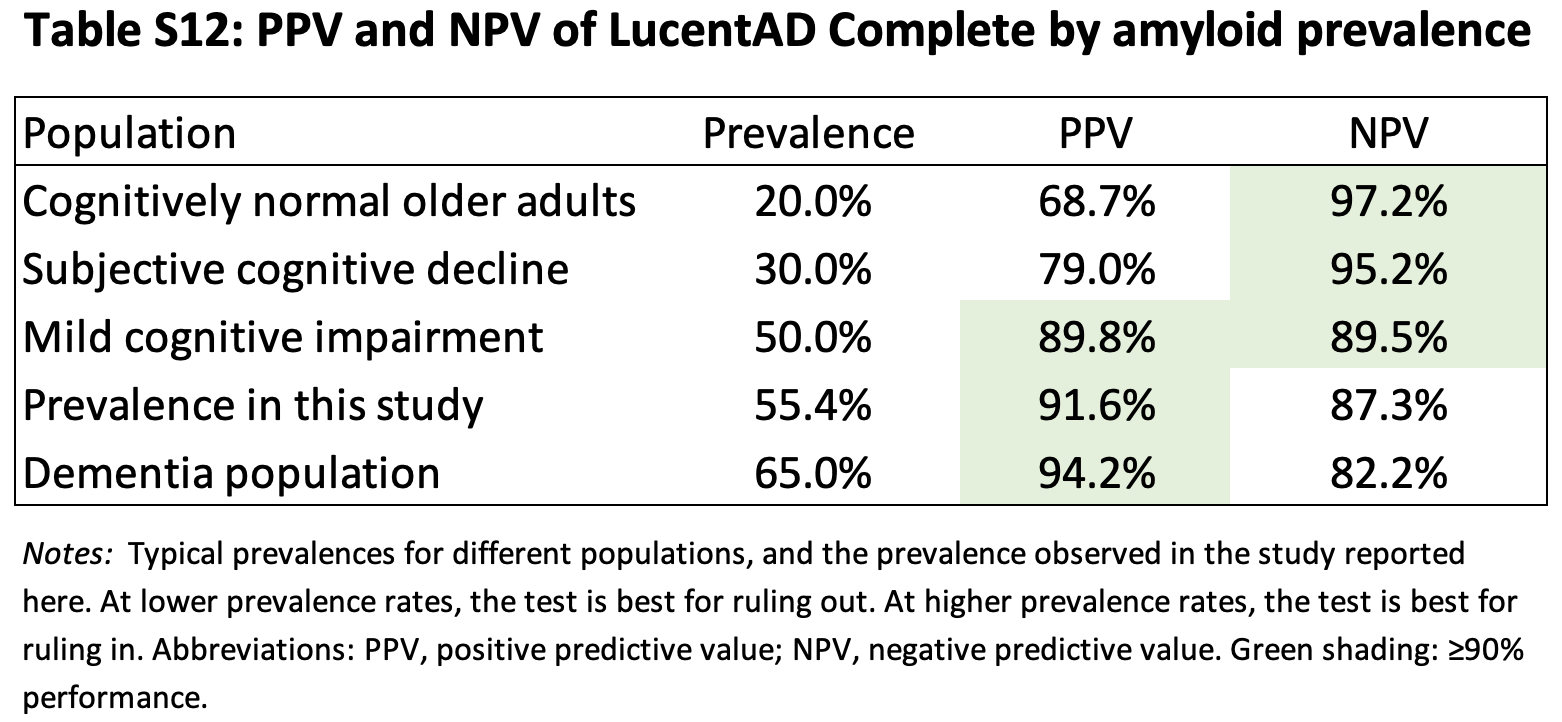
